# Impact of year-long cannabis use for medical symptoms on brain activation during cognitive processes

**DOI:** 10.1101/2024.04.29.24306516

**Authors:** Debbie Burdinski, Alisha Kodibagkar, Kevin Potter, Randi Schuster, A. Eden Evins, Satrajit Ghosh, Jodi Gilman

## Abstract

**Importance:** Cannabis is increasingly being used to treat medical symptoms, but the effects of cannabis use on brain function in those using cannabis for these symptoms is not known.

**Objective:** To test whether brain activation during working memory, reward, and inhibitory control tasks, areas of cognition impacted by cannabis, showed increases following one year of cannabis use for medical symptoms.

**Design:** This observational cohort study took place from July 2017 to July 2020 and is reported on in 2024.

**Setting:** Participants were from the greater Boston area.

**Participants:** Participants were recruited as part of a clinical trial based on seeking medical cannabis cards for anxiety, depression, pain, or sleep disorders, and were between 18 and 65 years. Exclusion criteria were daily cannabis use and cannabis use disorder at baseline.

**Main Outcomes and Measures:** Outcomes were whole brain functional activation during tasks involving working memory, reward and inhibitory control at baseline and after one year of cannabis use.

**Results:** Imaging was collected in participants before and one year after obtaining medical cannabis cards; 57 at baseline (38 female [66.7%]; mean [SD] age, 38.0 [14.6] years) at baseline, and 54 at one-year (37 female [68.5%]; mean [SD] age, 38.7 [14.3] years). Imaging was also collected in 32 healthy control participants (22 female [68.8%]; mean [SD] age, 33.8 [11.8] years) at baseline. In all groups and at both time points, functional imaging revealed canonical activations of the probed cognitive processes. No statistically significant difference in brain activation between the two timepoints (baseline and one-year) in those with medical cannabis cards and no association of changes in cannabis use frequency with brain activation were found.

**Conclusions and Relevance:** Findings suggest that adults do not show significant neural effects in the areas of cognition of working memory, reward, and inhibitory control after one year of cannabis use for medical symptoms. The results warrant further studies that probe effects of cannabis at higher doses, with greater frequency, in younger age groups, and with larger, more diverse cohorts.

Trial Registration:

NCT03224468, https://clinicaltrials.gov/

**Key Points:** *Question:* This study investigated the impact of year-long cannabis use for medical symptoms on brain activation during cognitive processes implicated in cannabis use.

*Findings:* Functional magnetic resonance imaging during a working memory, reward, and inhibitory control task was collected at baseline and after one year of medical cannabis card ownership. After one year, brain activation did not differ statistically from baseline and was not associated with changes in cannabis use frequency.

*Meaning:* The absence of activation differences suggests that adults using cannabis for medical conditions may not experience significant neural effects in regards to reward, working memory, or inhibitory control.

## Introduction

Accumulating evidence has shown that regular cannabis use can alter brain function, especially in networks that support working memory, cognitive control, and reward processing.^1^ Several prior reviews have described the functional impact of chronic cannabis use in both adults and adolescents,^2–4^ largely concluding that the domains of executive functioning and memory are most strongly affected by regular cannabis use.^5,6^ However, most of the evidence for brain changes with cannabis use is derived from between-group brain differences between those who use cannabis and those who do not, rather than from longitudinal changes at pre-and post-cannabis timepoints, raising the question of whether pre-existing differences between users and non-users underlie observed changes. Longitudinal studies, such as the ABCD study,^7^ are underway, however to date, few studies focused on adults using cannabis to treat medical symptoms. Little is known about effects of cannabis on the brain in medical populations, who may also experience illness-related cognitive weaknesses and may have different use patterns and age ranges compared to recreational users.

Delta-9-tetrahydrocannabinol (THC), the main psychoactive compound in cannabis, binds to endogenous cannabinoid CB1 receptors located in brain regions such as the hippocampus, amygdala, basal ganglia, prefrontal cortex, substantia nigra and globus pallidus,^8,9^ making frontal-limbic neurocircuitry particularly susceptible to cannabis-related effects in the brain.^10^ Specifically, THC binding inhibits release of neurotransmitters usually modulated through endocannabinoids.^11^ Many factors can modulate THC’s impact on the brain, including duration, frequency and quantity of use, age of initiation, potency, accompanying cannabidiol content, presence of cannabis use disorder (CUD), concurrent use of other substances, and sex and genetics.^12^

The question of how cannabis affects the brain is particularly relevant to those using cannabis to treat medical symptoms. Currently in the US, 37 states and the District of Columbia have medical cannabis programs, and enrollment in medical cannabis programs increased 4.5-fold from 2016 to 2020.^13^ In Massachusetts, obtaining a medical cannabis card (MCC) gives patients access to tax-exempt cannabis purchases and additional medical dispensaries. However, evidence for the effectiveness of plant-based cannabis for any medical condition is sparse.^14^ In dispensaries, a myriad of products (e.g. candies, gummies, smoked, vaped) are available to those using medically, and the neural effects of these products are unknown.

We sought to describe cognitive and brain-based changes in a longitudinal sample of participants beginning to use cannabis for medical symptoms, specifically for anxiety, depression, pain, and insomnia symptoms. We first conducted a pragmatic randomized clinical trial (RCT NCT03224468) of cannabis for medical symptoms to assess its effect on target symptoms when compared to a waitlist control group.^15,16^ In the current report, we describe the secondary outcomes of the clinical trial, a longitudinal analysis of task-based functional magnetic resonance imaging (fMRI) data from those who were assigned to obtain MCCs immediately. We explore the extent to which cannabis affects the brain during cognitive processes previously implicated in cannabis use, using neuroimaging tasks that probe working memory, reward processing, and inhibitory control. We hypothesized, based on previous literature,^17–19^ that a year of cannabis use would be associated with generally increased activation in brain regions underlying these processes and that an increase in cannabis use frequency would be associated with this increased activation, with few differences in task performance.

## Methods

### Study Recruitment

This study analyzed the fMRI data collected as part of a pragmatic, single-site, single-blind, RCT assessing MCC-seeking patients in the greater Boston area from July 1, 2017, to July 31, 2020 (see *NCT03224468*).^15,16^ Briefly, participants were between the ages of 18 and 65, and were seeking to obtain MCCs for the first time for depression, anxiety, pain, or insomnia symptoms, the most commonly reported conditions in those seeking cannabis for symptom management. Exclusion criteria included daily cannabis use, CUD diagnosis at screening or baseline, cancer, psychosis, and current substance use disorders (except for mild or moderate alcohol use disorder and nicotine use disorder).

### Study Protocol

Participants were randomized to either receive their MCC immediately or to delay acquisition by 12 weeks. In the current study, only data from the immediate acquisition MCC group as well as an age-and sex-matched healthy control (HC) group are presented. Demographic variables were collected at baseline using self-report. Behavioral data collected at baseline, 2, 4, 12, 24, and 52 weeks included past-month frequency of cannabis use and validated scales to assess CUD, insomnia, depression and anxiety, and pain symptoms. In addition, urinalysis for cannabis metabolites was conducted at the time of study visits. Structural and functional brain imaging data was collected only in the MCC group (n=70, eFigure 1). At baseline imaging was also collected in the HC group (n=32). fMRI tasks included a working memory (N-back) task, a reward processing (monetary incentive delay, MID) task and an inhibitory response (stop signal, SST) task.^20–22^ See the supplementary methods for a detailed description of the experimental paradigm (eFigures 2-4). Participants provided written informed consent and were financially compensated for their participation in the study. The clinical trial was approved by the Massachusetts General Brigham institutional review board. This report focuses on the analysis of the task-based functional imaging, which followed a pretest-posttest design with a control group at baseline. Clinical outcomes of the RCT are reported elsewhere.^15,16^

**Figure 1:**
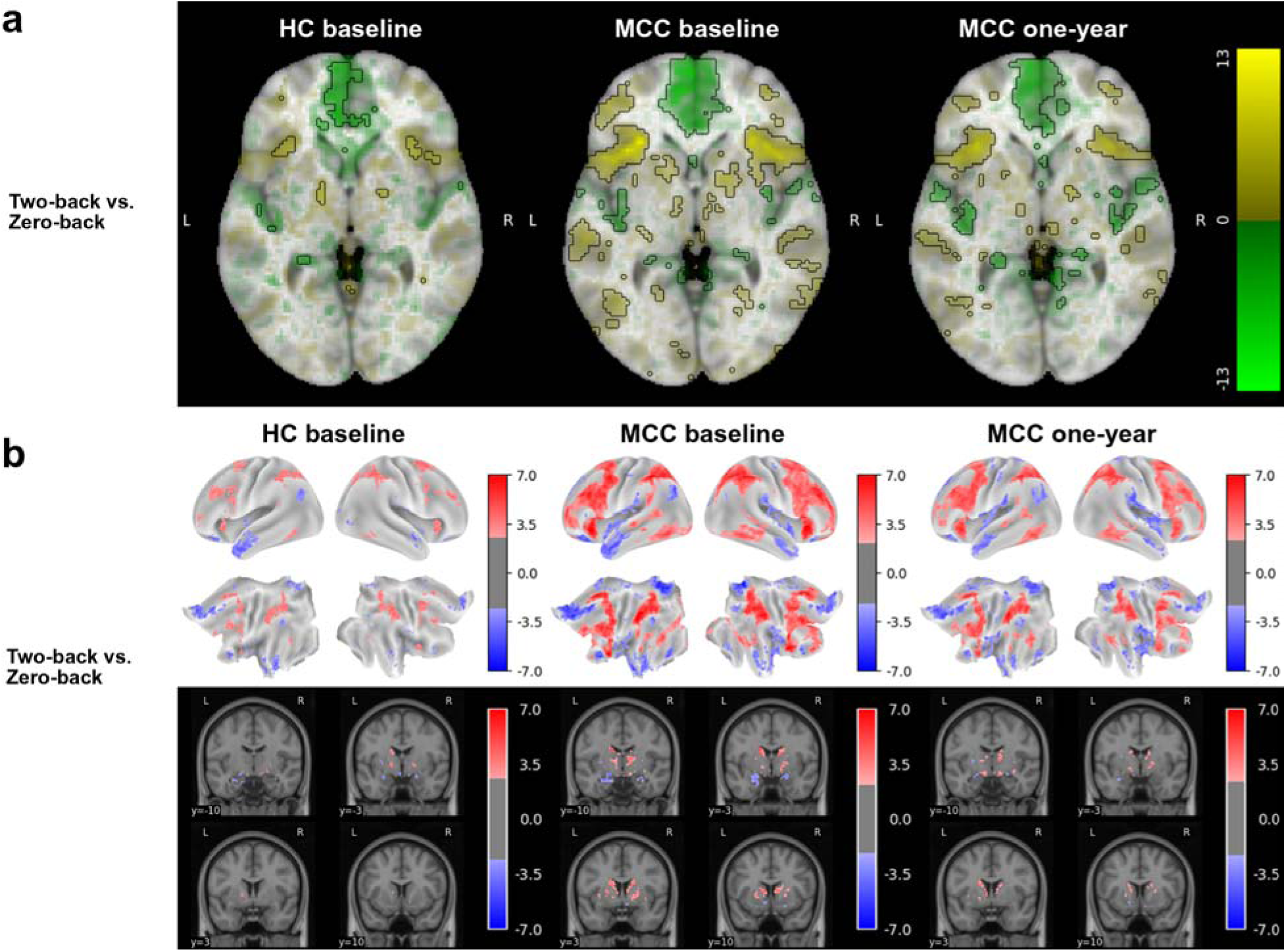
Brain activation for the N-back task’s two-back vs. zero-back contrast across groups and timepoints The HC group at baseline (n=22), the MCC group at baseline (n=40), and the MCC group at one-year (n=40) did not show activation differences between the two groups at baseline or between the two time points of the MCC group. Cannabis use frequency changes were not associated with brain activation at one-year. Figure 1a: Voxel-wise average brain activation, colored by effect size and opacity-scaled by z-scores with significance threshold (FDR=0.05) outlined, for the two-back vs. zero-back contrast of the N-back task. z-thresholds: HC group at baseline=3.05; MCC group at baseline=2.47, and MCC at one-year=2.60. Colorbar displays effect size. Figure 1b: Grayordinate-wise average brain activation z-scores above significance threshold (FDR=0.05) for the two-back vs. zero-back contrast of the N-back task. z-thresholds: HC group at baseline=2.60), MCC group at baseline=2.21, and MCC at one-year=2.35. Colorbar displays z-scores. HC=healthy control; MCC=medical cannabis card.

**Figure 2:**
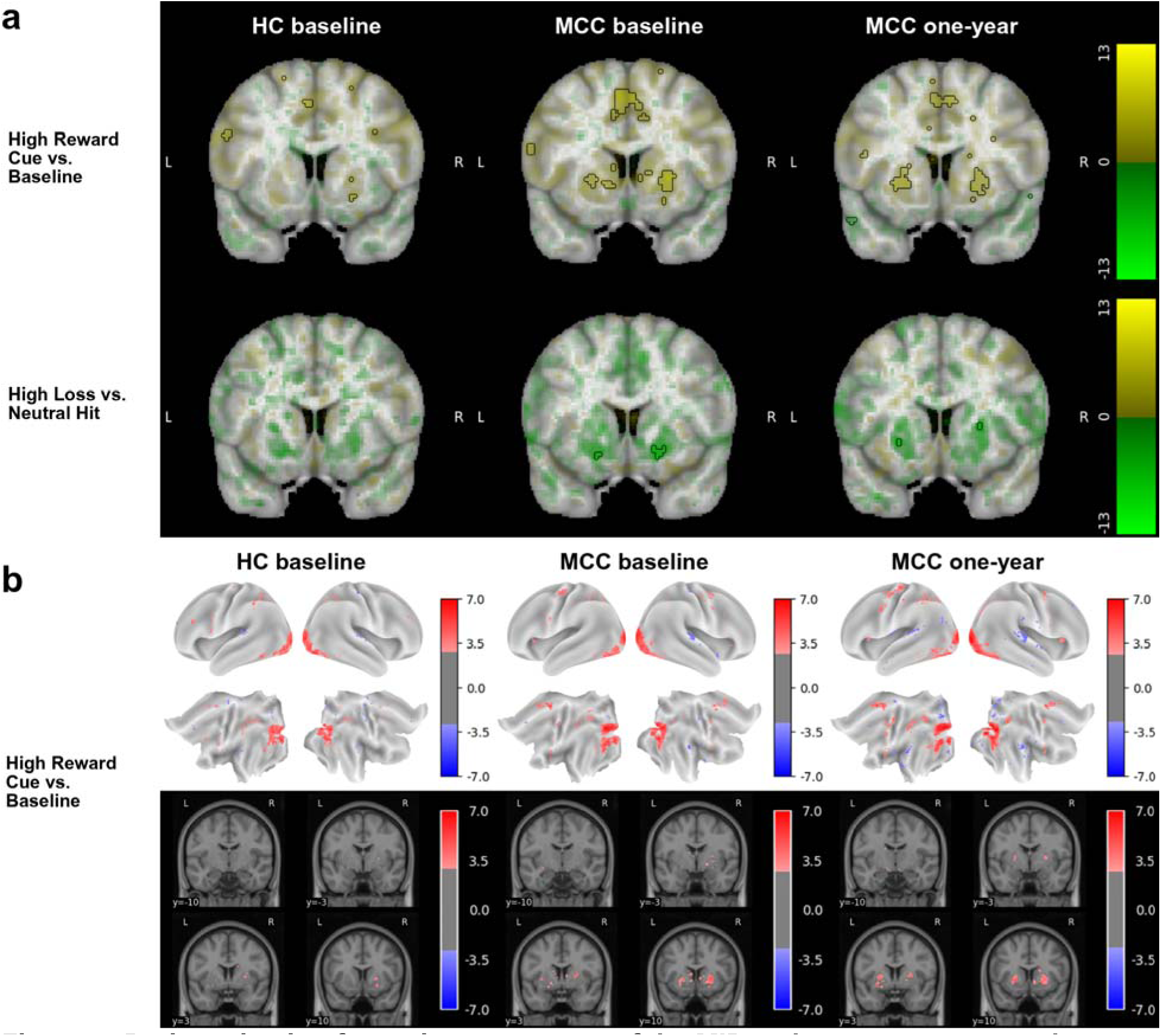
Brain activation for various contrasts of the MID task across groups and timepoints The HC group at baseline (n=23), the MCC group at baseline (n=35), and the MCC group at one-year (n=40; except High Loss vs. Neutral Hit: n=39) did not show activation differences between the two groups at baseline or between the two time points of the MCC group. Cannabis use frequency changes were not associated with brain activation at one-year. Figure 2a: Voxel-wise average brain activation, colored by effect size and opacity-scaled by z-scores with significance threshold (FDR=0.05) outlined, for the high reward cue vs. baseline contrast (top row) and the high loss vs. neutral hit feedback contrast (bottom row) of the MID task. z-thresholds: HC group at baseline=3.37 (top) and undetermined (bottom), MCC group at baseline=3.24 (top) and 4.34 (bottom), and MCC at one-year=3.10 (top) and 4.65 (bottom). Note that activation for the other contrasts was below threshold. Colorbar displays effect size. Undetermined z-threshold indicates that no voxel was statistically significant. Figure 2b: Grayordinate-wise average brain activation z-scores above significance threshold (FDR=0.05) for the high reward cue vs. baseline contrast of the MID task. z-thresholds: HC group at baseline=2.91, MCC group at baseline=2.74, and MCC at one-year=2.70. Note that activation for the other contrasts was below threshold. Colorbar displays z-scores. HC=healthy control; MCC=medical cannabis card.

### Participant Characteristics Analysis

Demographic metrics were compared across the four groups (HC participants at baseline, MCC participants at baseline, MCC participants at one-year, MCC participants imaged at both time points) using a one-way ANOVA for numerical variables, a Fisher’s exact test for categorical variables with less than 5 observations in a category, and a Chi-squared test for all other categorical variables. Cannabis metrics in the MCC participants who were imaged at both time points were compared across the two time points (baseline and one-year) using a t-test for numerical variables, a Fisher’s exact test for categorical variables with less than 5 observations in a category, and a Chi-squared test for all other categorical variables.

### MRI Data Analysis

#### Pre-Processing

Volumes were pre-processed using version 23.0.1 of the *fMRIPrep* software, which included head-motion estimation, slice time correction, fieldmap-based distortion correction, EPI to T1 registration and resampling to both MNI volumetric and grayordinate space.^23,24^ Quality control metrics were derived using the *MRIQC* software.^25^ See the supplementary methods for details on MRI acquisition and pre-processing.

#### General Linear Modeling

To corroborate the robustness of our findings, two general linear model analyses were conducted, one in volumetric and the other in grayordinate space, in *Python 3.9.13* using the package *Nilearn 0.9.2*.^26,27^ Time series were scaled and smoothed. First-level modeling removed noise using motion, drift and anatomical CompCor regressors. For the N-back task, a single image contrast of the two-back vs. zero-back stimuli was calculated. For the MID task, given the number of possible contrasts, this study focused on those from two recent publications.^28,29^ Anticipation contrasts included high reward vs. neutral anticipation, low reward vs. neutral anticipation, reward vs. neutral anticipation, high reward vs. low reward anticipation, high reward vs. implicit baseline, high loss vs. neutral anticipation, low loss vs. neutral anticipation, and high loss vs. low loss anticipation. Feedback contrasts included high reward vs. neutral hit feedback, reward vs. missed reward feedback, high loss vs. neutral hit feedback and loss vs. avoided loss feedback. For the SST task, contrasts calculated include correct inhibition (successful STOP vs. GO), incorrect inhibition (unsuccessful STOP vs. GO), and unsuccessful inhibitory control (unsuccessful STOP vs. successful STOP). Two runs were collected for the MID and SST tasks, which were combined using a second linear model prior to group-modeling.

Individual effect sizes for the contrasts were passed to a group-level model to assess group averages at a given time point, differences across groups and time points, and the role of cannabis use frequency (two-sided tests for contrast or parameter significance in the setting of an ordinary least squares regression model with covariates). Covariates included sex, age, and past-month cannabis use frequency (at the time point of the analysis unless specified below), mean-centered for numerical variables.

Whole brain activation was compared between MCC users and a matched control group at baseline using a contrast between group-level intercepts. For the across time analyses of the MCC group, observations were limited to those participants with imaging at both timepoints. It was assessed whether, on average, there was a whole brain activation difference between baseline and one-year timepoints in an individual, controlling for baseline cannabis use frequency to account for individual differences in use at the outset of the study. Further, the effect of the change in cannabis use frequency across time on whole brain activation at one-year was assessed, adding the additional covariates of brain activation and cannabis use frequency at baseline.

Runs were excluded if they were statistical outliers at a study time point based on their signal-to-noise ratio, temporal signal-to-noise ratio, ghost-to-signal ratio along the two phase encoding directions, if their mean framewise displacement (FD) was above 0.2 (unless <30% of a scan had a FD above the FD threshold), or if more than 30% of their volumes were motion outliers as determined by *fMRIPrep*. Further FD cutoffs were assessed to corroborate our findings. Effect sizes at the group level were standardized for visualization purposes.^30,31^ Statistical inference was FDR controlled at 0.05 using the Benjamini-Hochberg procedure. See the supplementary methods for details on the general linear modeling approach.

### Behavioral Data Analysis

Differences between the HC and MCC groups at baseline and between the two time points of the MCC participants in regards to reaction time and accuracy across the two-back, zero-back and combined stimuli of the N-back task were assessed using a two-sided t-test (paired in the case of MCC participants across time). Differences between the groups (HC at baseline, MCC at baseline, MCC at one-year) for stop signal reaction time (the inferred mean latency between the stop signal and response inhibition; SSRT) were estimated via an additive multilevel linear model with a participant-varying intercept.

## Results

### Participant Characteristics

We collected brain imaging data in 70 MCC and in 32 control participants. Briefly, at baseline 57 MCC participants (38 female [66.7%]; mean [SD] age, 38.0 [14.6] years) and 32 control participants (22 female [68.8%]; mean [SD] age, 33.8 [11.8] years) were imaged. After one year 54 MCC participants (37 female [68.5%]; mean [SD] age, 38.7 [14.3] years) were imaged. Out of all MCC participants, 41 presented for imaging at both time points (28 female [68.3%]; mean [SD] age, 39.6 [14.4] years). The four groups did not differ significantly in any of the characteristics assessed, including sex, age, race, ethnicity, education years, and handedness, and the three MCC subsets did not differ in the symptoms they enrolled in the study for (Table 1).

**Table 1:** Characteristics of the study participants. ^a^ HC baseline corresponds to the imaging control group at baseline ^b^ MCC baseline corresponds to the medical cannabis card group’s participants imaged at baseline ^c^ MCC one-year corresponds to the medical cannabis card group’s participants imaged at one-year ^d^ MCC paired corresponds to the medical cannabis card group’s participants imaged at both timepoints

| Items | Levels | HC baseline<sup>a</sup> | MCC baseline<sup>b</sup> | MCC one-year<sup>c</sup> | MCC paired<sup>d</sup> |
| --- | --- | --- | --- | --- | --- |
| n | NA | 32 | 57 | 54 | 41 |
| Sex, n (%) | Female | 22 (68.8) | 38 (66.7) | 37 (68.5) | 28 (68.3) |
| Age, mean (SD) | NA | 33.8 (11.8) | 38.0 (14.6) | 38.7 (14.3) | 39.6 (14.4) |
| Race, n (%) | Black | 2 (6.2) | 6 (10.5) | 4 (7.4) | 2 (4.9) |
| Race, n (%) | Other | 3 (9.4) | 6 (10.5) | 2 (3.7) | 2 (4.9) |
| Race, n (%) | White | 27 (84.4) | 45 (78.9) | 48 (88.9) | 37 (90.2) |
| Ethnicity, n (%) | Hispanic or Latino | 3 (9.4) | 1 (1.8) | 1 (1.9) | 1 (2.4) |
| Education years, mean (SD) | NA | 17.8 (2.7) | 16.6 (2.4) | 17.0 (2.3) | 17.1 (2.3) |
| Condition, n (%) | Depression/anxiety symptoms | 0 | 27 (47.4) | 24 (44.4) | 19 (46.3) |
| Condition, n (%) | Insomnia symptoms | 0 | 12 (21.1) | 13 (24.1) | 9 (22.0) |
| Condition, n (%) | Pain symptoms | 0 | 18 (31.6) | 17 (31.5) | 13 (31.7) |
| Handedness, n (%) | Right-handed | 29 (90.6) | 49 (86.0) | 46 (85.2) | 35 (85.4) |

To capture changes in cannabis use throughout the year of having the MCC, severity of CUD symptoms, CUDIT-R summed score, cannabis use frequency per month, and positive urine THC were compared between the baseline and one-year time point (eTable 1). There was a statistically significant difference between the two time points in all metrics, which all tended towards higher cannabis use at the one-year time point compared to at baseline.

### N-back Task

Activation in prefrontal and parietal cortical regions was observed for the two-back vs. zero-back image contrast in all three participant groups, including MCC participants at baseline and one-year as well as control participants (Figure 1). No difference in activation between the groups at baseline (HC n=22, MCC n=40) or between the two timepoints of the MCC group (n=25), and no effect of cannabis use frequency change on one-year activation of the MCC group were noted at the statistical significance threshold.

Control participants compared to MCC participants at baseline, as well as MCC participants compared across the two time points restricted to those with imaging at both time points, also did not differ in accuracy or reaction time for the zero-back or two-back conditions of the N-back task at a statistical threshold of p<0.05 (eTable 2).

### MID Task

During all cue contrasts, activation in the bilateral basal ganglia was observable in all three participant groups, though only reached statistical significance in the High Reward Cue vs. Baseline contrast (Figure 2, eFigures 5,6).^32^ During the reward vs. missed reward feedback contrast, activation in the bilateral basal ganglia is observed, while during all other feedback contrasts, deactivation in the bilateral basal ganglia and insula is observed, though only statistically significant in the high loss vs. neutral hit contrast in the volumetric analysis (Figure 2, eFigures 5,6). No difference in activation between the groups at baseline (HC n=23, MCC n=35) or between the two timepoints of the MCC group (n = 21 for High Reward vs. Neutral Hit and High Loss vs. Neutral Hit contrasts; n=22 for all other contrasts), and no effect of cannabis use frequency change on one-year activation of the MCC group were noted at the statistical significance threshold.

### SST Task

Activation in inhibitory control-related regions, including the right inferior frontal gyrus, frontal gyrus, and insula, was observed for the correct inhibition, incorrect inhibition and successful inhibitory control contrasts in all three participant groups, including MCC participants at baseline and one-year as well as control participants (Figure 3, eFigures 7,8). No difference in activation between the groups at baseline (HC n=25, MCC n=40) or between the two timepoints of the MCC group (n=26), and no effect of cannabis use frequency change on one-year activation of the MCC group were noted at the statistical significance threshold.

**Figure 3:**
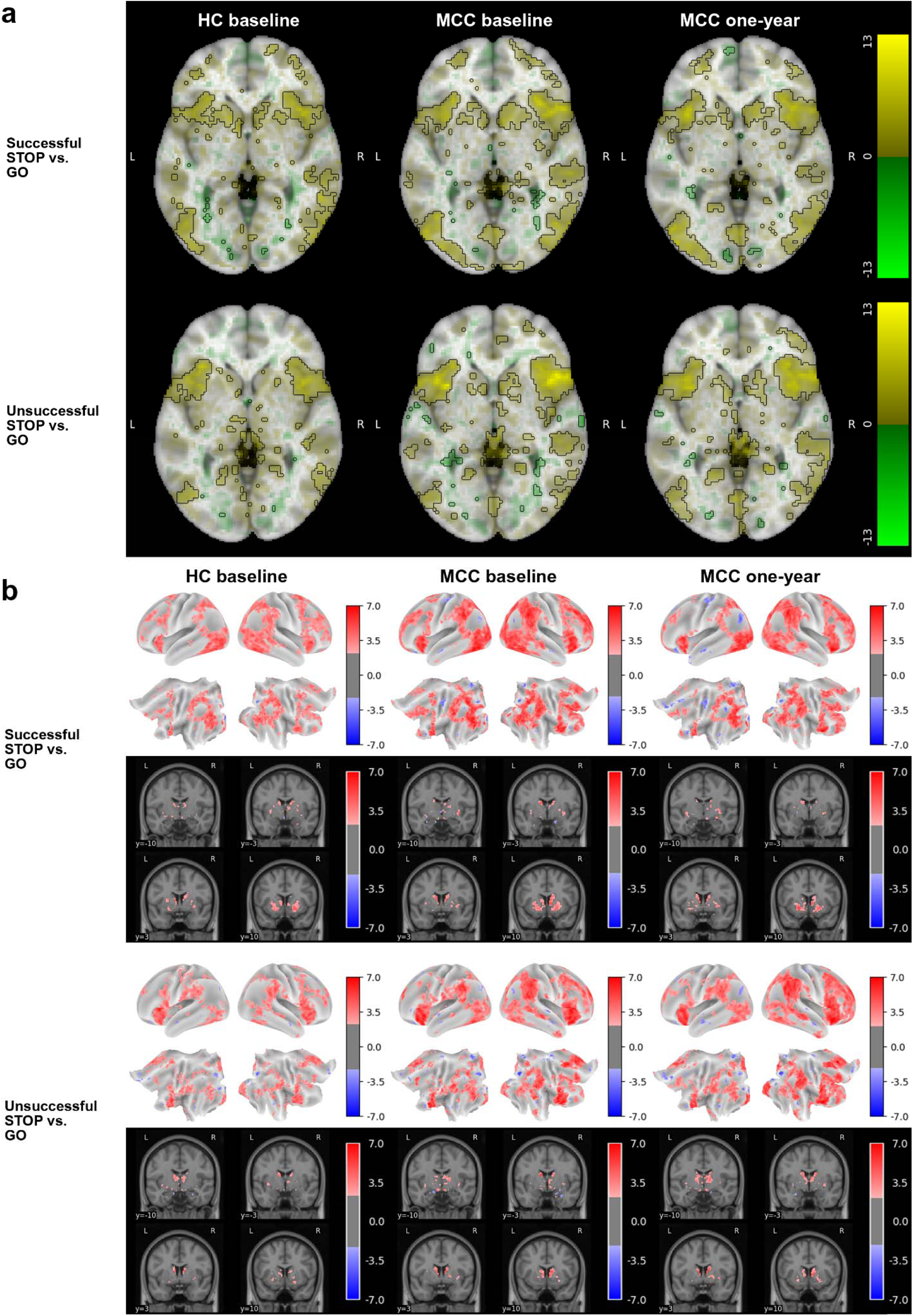
Brain activation for the SST task’s two STOP vs. GO contrasts across groups and timepoints The HC group at baseline (n=25), the MCC group at baseline (n=40), and the MCC group at one-year (n=44) did not show activation differences between the two groups at baseline or between the two time points of the MCC group. Cannabis use frequency changes were not associated with brain activation at one-year. Figure 3a: Voxel-wise average brain activation, colored by effect size and opacity-scaled by z-scores with significance threshold (FDR=0.05) outlined, for the successful STOP vs. GO contrast (top row) and the unsuccessful STOP vs. GO contrast (bottom row) for the SST task. z-thresholds: HC group at baseline=2.77 (top) and 2.87 (bottom); MCC group at baseline=2.56 (top) and 2.62 (bottom), and MCC at one-year=2.62 (top) and 2.55 (bottom). Colorbar displays effect size. Figure 3b: Grayordinate-wise average brain activation z-scores above significance threshold (FDR=0.05) for the successful STOP vs. GO contrast (top row) and the unsuccessful STOP vs. GO contrast (bottom row) for the SST task. z-thresholds: HC group at baseline=2.25 (top) and 2.31 (bottom); MCC group at baseline=2.15 (top) and 2.20 (bottom), and MCC at one-year=2.22 (top) and 2.13 (bottom). Colorbar displays z-scores. HC=healthy control; MCC=medical cannabis card.

At baseline, HC participants had significantly faster SSRT (259土43ms) compared to MCC participants (276土40ms), β=-16.2 (SE=7.7), p = 0.035. MCC participants had a nonsignificant reduction in SSRT from baseline (276土40ms) to year 1 (264土43ms), β =-10.0 (SE=5.1), p = 0.050.

Relaxing the FD threshold from 0.2 to 0.3 and not removing any quality control outliers did not change the results above (eFigures 9-14).

## Discussion

After year-long cannabis use for medical symptoms, we did not observe functional differences during working memory, reward processing, or inhibitory control tasks or an association of changes in cannabis use frequency with brain activation. Similarly, no significant changes in behavioral performance emerged. This suggests that cannabis use for medical purposes, within the snapshot of cognition captured by these tasks and within a mostly older, white, female and generally well-educated population, does not have a significant neural impact.

Prior studies have found that cannabis use, especially in adolescents, is associated with impairments in cognitive processes beyond acute intoxication.^11^ Such studies have largely been cross-sectional, have generally not focused on adults using cannabis for medical purposes, and have focused primarily on heavy cannabis use. Further, conclusions of studies comparing individuals who use cannabis with those who do not often are limited by inherent group differences at the outset. Thus, the neural impact of cannabis use for medical purposes has remained largely unknown. Because participants in this study did not use cannabis heavily at baseline and obtained MCCs for the first time for their medical symptoms, the study was uniquely positioned to examine the brain before and after adults began to use cannabis regularly, a period that is difficult to capture. This study is one of the first studies to evaluate brain activation differences in an ecologically valid setting in those who began using cannabis for medical and psychological symptoms.

Memory is one of the most consistently reported processes that is affected by cannabis.^33^ Prior studies comparing those who use cannabis to those who do not have found significant changes in activation of frontal regions during the N-back task,^34–37^ though it should be noted that other studies did not report statistically significant difference in any regions.^38,39^ Reward-related activation has also been implicated in cannabis use, as prior studies have found significant changes in activation of striatal regions during the MID task.^40–45^ Finally, inhibitory control activation differences have been reported, ^46,47^ particularly in fronto-basal-ganglia circuits.^48^ However, these studies mainly consist of those who began using cannabis during adolescence or those who use cannabis frequently. Contrary to our initial hypothesis and this literature, our findings indicate that activation to working memory, reward processing, and inhibitory control tasks is largely unchanged in adults using cannabis to alleviate medical symptoms for one year. There are potential reasons for the lack of neural changes in this study compared to previous studies of recreational cannabis, including differences in use patterns, amounts, motivations for use, lower use in adolescence or other factors.

The findings of the study are corroborated by the results of the control group and by the brain activation patterns observed at both timepoints. Brain activation differences were not found between healthy controls and MCC participants at baseline, suggesting that the MCC group did not differ significantly at baseline from those who were not intending to use cannabis, thus addressing some of the limitations in previous studies. The working memory and inhibitory control tasks yielded statistically significant canonical activations in the control and MCC group at baseline and in the MCC group at the one-year timepoint.^20,28,48,49^ The reward processing task also yielded canonical activations for the cue contrasts.^28,29,32,50,51^ Activations to the feedback contrasts were also in line with what has been observed in the past, though the literature is less consistent in regards to the feedback contrast.^29,50,51^ Of note, only two of the MID contrasts reached statistical significance. This suggests that the response to the task was more heterogeneous in this participant sample, and perhaps a larger sample or a differently designed reward task would have been needed to achieve more robust activation. It was recently noted that the MID task has low reliability, with the most consistent contrasts being those involving an implicit baseline, which is consistent with our limited findings.^52^ Overall, the activation patterns of all three tasks were consistent with those observed in the literature, demonstrating that in the comparison across time, brain activation from the desired cognitive processes is being compared.

### Limitations

This study should be interpreted in light of its limitations. First, the sample was predominantly female, white, older, and well-educated, which may limit the generalizability of our findings. Future studies should thus focus on recruiting a more diverse sample. Second, longitudinal effects not related to cannabis use, such as age-related changes, cannot be separated from cannabis-specific effects. Thus, future studies should compare brain imaging from a non-using control group across two time points as well. It is further possible that the lack of difference in task-based activation was due to limited power. We note that the maximal absolute effect size difference between the two time points in the MCC group is smaller than the maximal absolute effect size across the two time points for all three tasks. Thus, even if we did not detect a difference despite one existing, the change in brain activation after year-long cannabis use would still be small.

Importantly, adult-onset use of cannabis for medical symptoms may have different brain implications compared to adolescent use, something that should be studied in future investigations. Moreover, comorbid conditions (eg. depression or pain) may influence the impact of cannabis on the brain. While our sample size was too small for a subgroup analysis of each of the symptoms that participants sought the MCC for, future studies should enroll sufficient participants to be able to discover potential differences of the impact of cannabis across symptoms. Lastly, in this study participants freely chose cannabis products at local dispensaries to simulate the typical use of cannabis for medical symptoms. It is important to note that since participants could choose their own products and doses, it is possible that doses of cannabinoids were too low to observe brain changes. Further research is warranted to understand how differences in product type, amounts, and patterns of use might affect the brain in cannabis users for medical symptoms.

The tasks included in this study only capture a snapshot of cognition, probing aspects of working memory, reward processing and inhibitory control. Brain activation during tasks that probe the same cognitive processes from a different angle or that probe other cognitive processes could change after yearlong cannabis use for medical symptoms. While this study selected classic tasks probing areas of cognition known to be affected by recreational cannabis use, it cannot ascertain that no facet of cognition is impacted by medical use of cannabis and future studies should explore additional tasks or functional connectivity independent of a specific task.

### Conclusions

In this study, brain activation during working memory, reward processing, and inhibitory control tasks was not statistically different after year-long cannabis use for medical symptoms and no effect of changes in cannabis use was noted. Our results suggest that adults who use cannabis, generally with light-to-moderate patterns, for such symptoms experience no significant long-term neural effects in these areas of cognition.

## Data Availability

The data are shared with and accessible through the ENIGMA consortium. All preprocessing and analysis code is available on GitHub.

https://github.com/burdinskid13/cannabis-paper

## Author Contributions

Drs Gilman and Ghosh had full access to all the data in the study and take responsibility for the integrity of the data and the accuracy of the data analysis.

*Concept and design:*Burdinski, Gilman, Ghosh, Evins.

*Acquisition, analysis, or interpretation of data:* Burdinski, Kodibagkar, Potter, Gilman, Ghosh.

*Drafting of the manuscript:* Burdinski, Kodibagkar, Gilman.

*Critical revision of the manuscript:* for important intellectual content Burdinski, Gilman, Ghosh, Evins, Schuster.

*Statistical analysis*: Burdinski, Potter.

*Obtained funding:* Gilman, Ghosh.

*Administrative, technical, or material support:* Burdinski, Gilman, Ghosh. Supervision: Gilman, Ghosh.

## Meeting Presentation

This work was presented at the 2023 Organization of Human Brain Mapping Meeting; July 22-26, 2023; Montréal, Canada.

## Data Sharing Statement

The data are shared with and accessible through the ENIGMA consortium.^53^ All preprocessing and analysis code is available here: https://github.com/burdinskid13/cannabis-paper.

