## Supplement 1 for "Impact of year-long cannabis use for medical symptoms on brain activation during cognitive processes"

### Supplementary Content

**Supplementary Content** 1
**Supplementary Methods** 3
Study protocol 3
eFigure 1: Study flow diagram 3
Experimental Paradigm 3
Behavioral Data 3
N-Back Task 3
eFigure 2: Schematic of the N-back task 4
MID Task 4
eFigure 3: Schematic of the MID task 5
SST Task 5
eFigure 4: Schematic of the SST task 6
MRI Data Analysis 6
MRI Acquisition 6
Pre-Processing 7
First-Level Analysis 8
Group-Level Analysis 9
**Supplementary Results** 10
Cannabis Metrics 10
eTable 1: Statistically significant difference in cannabis-related characteristics after one year of cannabis use for medical symptoms 10
N-back Accuracy and Reaction Time 11
eTable 2: No statistically significant difference in N-back accuracy and reaction time compared to control or across time 11
Results From Additional Contrasts 12
eFigure 5: Brain activation for additional contrasts of the MID task across groups and timepoints from the volumetric analysis 14
eFigure 6: Brain activation for additional contrasts of the MID task across groups and timepoints from the grayordinate analysis 14
eFigure 7: Brain activation for additional contrasts of the SST task across groups and timepoints from the volumetric analysis 15
eFigure 8: Brain activation for additional contrasts of the SST task across groups and timepoints from the grayordinate analysis 15
Main Results With Varying Outlier Removal 16
eFigure 9: Brain activation for the N-back task's two-back vs. zero-back contrast across groups and timepoints with no outliers removed 16
eFigure 10: Brain activation for the N-back task's two-back vs. zero-back contrast across groups and timepoints with the FD cutoff relaxed to 0.3 17
eFigure 11: Brain activation for various contrasts of the MID task across groups and timepoints with no outliers removed 19
eFigure 12: Brain activation for various contrasts of the MID task across groups and timepoints with the FD cutoff relaxed to 0.3 21
eFigure 13: Brain activation for the SST task's two STOP vs. GO contrasts across groups and

timepoints with no outliers removed 23
eFigure 14: Brain activation for the SST task's two STOP vs. GO contrasts across groups and timepoints with the FD cutoff relaxed to 0.3 25
**Supplementary References** **26**

#### Supplementary Methods

##### Study protocol

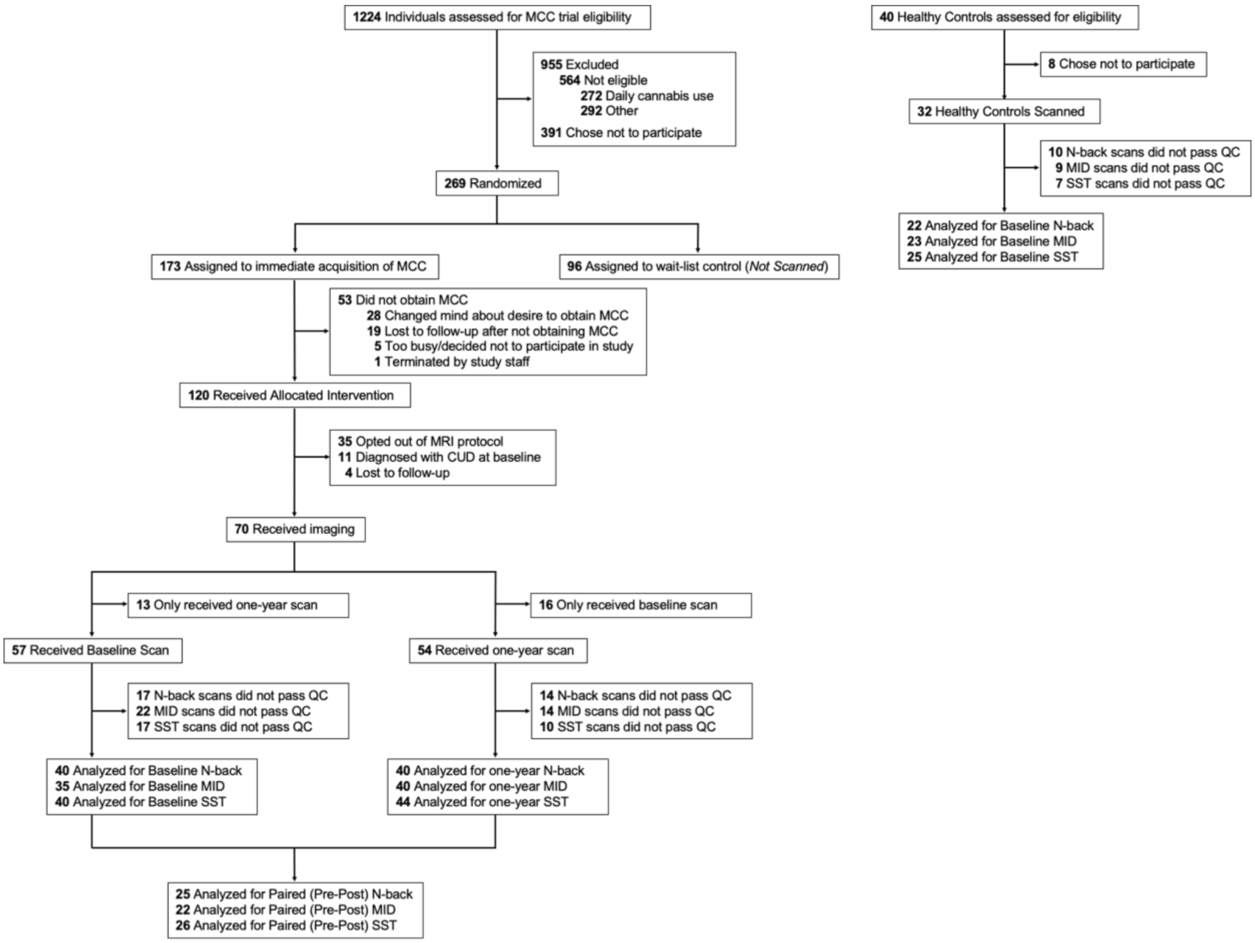


**eFigure 1: Study flow diagram**

The study used data that was collected as part of a larger clinical trial of cannabis for medical symptoms. Imaging data was collected in one arm of the trial and in a control group.

##### Experimental Paradigm

###### Behavioral Data

Frequency of cannabis use was assessed using a likert-scale of use throughout the past month (Less than once a week; Less than once a month; 1-2 days a week; 3-4 days a week; 5-6 days a week; Once or more per day). Cannabis use disorder (CUD), insomnia, depression and anxiety, and pain symptoms were assessed using the Cannabis Use Disorder Identification Test Revised (CUDIT - R), the Athens Insomnia Scale (AIS), the Hospital Anxiety and Depression Scale (HADS), and the Brief Pain Inventory (BPI), respectively.

#### N-Back Task

In the N-back task, participants were asked to press a button either on the same letter as two letters back (two-back) or on a predetermined letter (zero-back). There were 3 two-back blocks and 3 zero-back blocks, which were presented in an alternating fashion in a single run, starting with a zero-back block. After zero-back blocks there was a 2000ms gap, while after two-back blocks there was a 15000ms rest period. Participants were shown a different pseudorandomized letter sequence in each block and each sequence had 29 letters per round, of which 5 required a button press. Letters were presented for 500ms, followed by a 1500ms blank interstimulus screen and participants could respond at any time during this 2000ms window. Accuracy and reaction time were calculated for the zero-back and two-back trials individually and for the whole task. See eFigure 2 for a graphical overview.

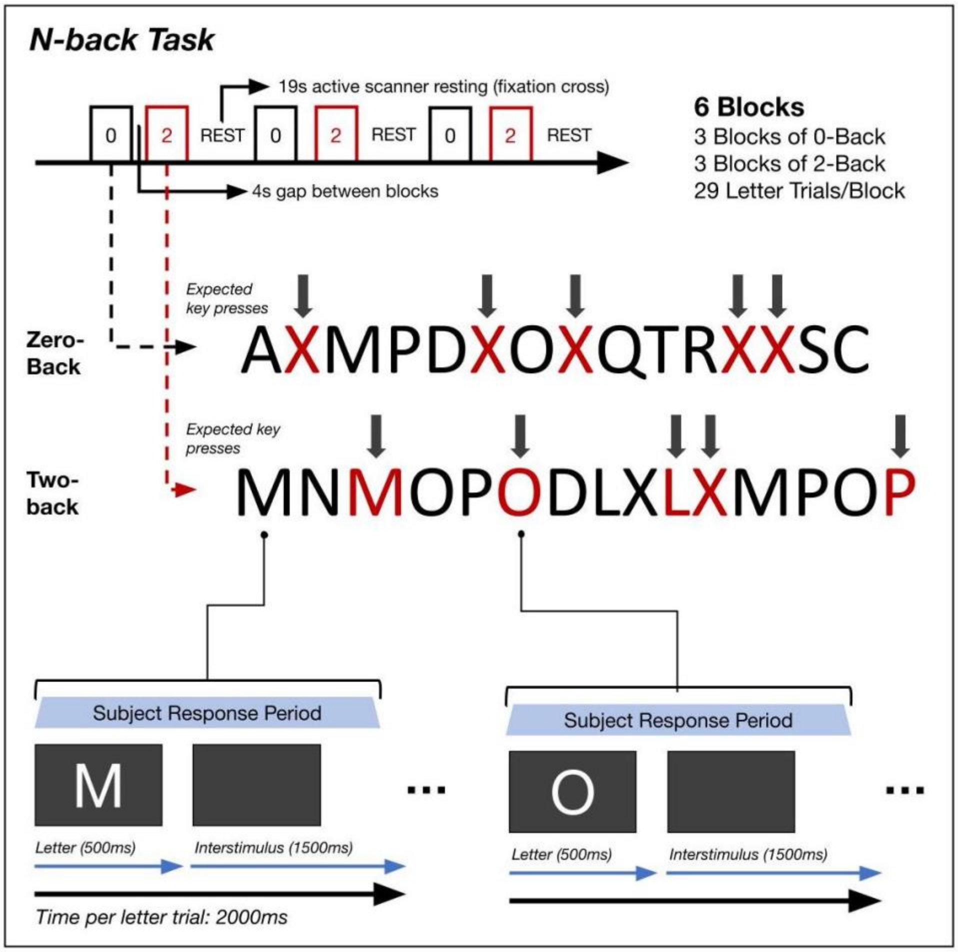


**eFigure 2: Schematic of the N-back task**

The N-back task consists of one run with 6 blocks of alternating zero-back and two-back blocks. Participants press a button either on the same letter as two letters back (two-back) or on a predefined letter (zero-back) within a time frame of 2000ms.

#### MID Task

In the MID task, participants were asked to press a button as fast as possible following a fixation cross of variable length to either win or avoid losing a certain monetary amount. The task consisted of 2 runs with 50 trials each (10 $5 win, 10 $5 loss, 10 $0.20 win, 10 $0.20 loss, 10 neutral). Each trial started with the presentation of the cue slide (2000ms) showing an outcome (win/loss/neutral), followed by a fixation cross lasting a randomized duration (1500-

4000ms). Then the participant was given a certain period of time to respond. The response period was initially set at a participant's reaction time and recalculated every 3 trials based on the participant's overall accuracy (between 150ms and 500ms). After each run a score was calculated based on wins and losses and the combined score was paid out to the participant at the end of the second run. See eFigure 3 for a graphical overview.

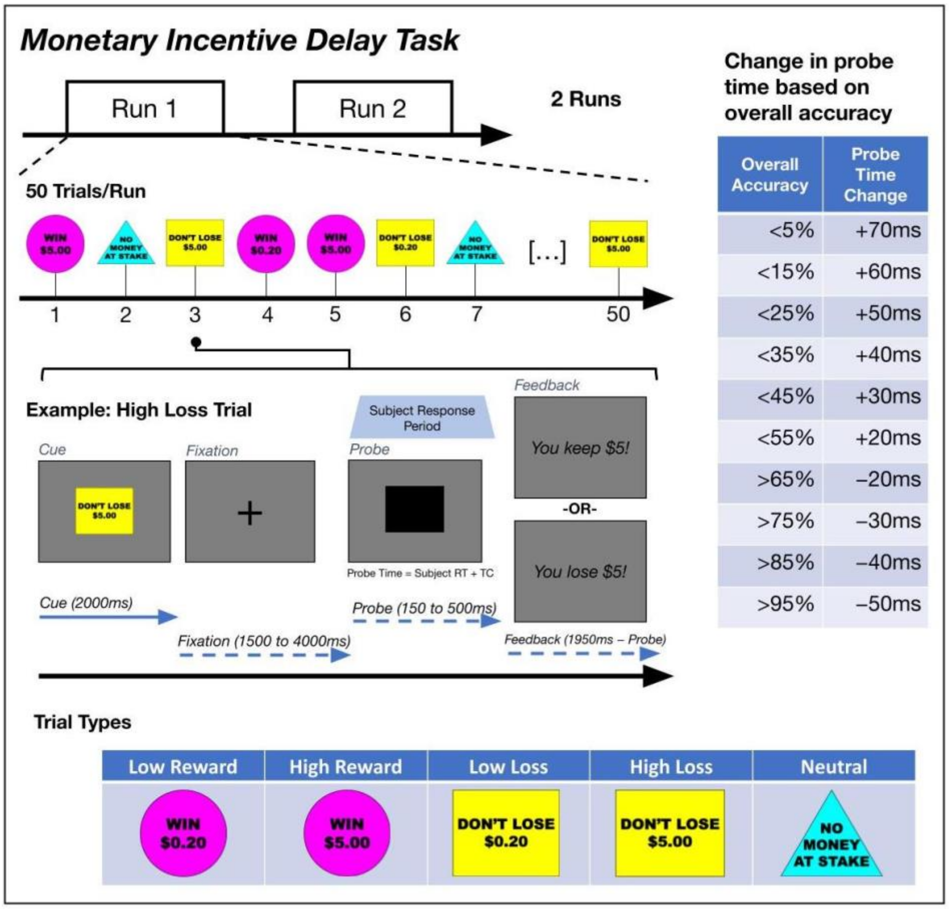


#### eFigure 3: Schematic of the MID task

The MID task consists of two runs in which participants are presented cues to either win or lose a high or low amount of money. Then they have to press a button as fast as possible following a fixation cross of variable length and receive feedback on whether they gain or lose money. The response period is altered based on performance and the monetary reward is paid out to participants at the end of the two runs.

##### SST Task

In the SST task, participants were asked to press a button upon presentation of an arrow (GO trial) unless it was immediately followed by a stop signal (STOP trial). The task consisted of 2 runs with 104 trials each (78 GO/ 26 STOP). A GO trial consisted of a 200ms fixation cross followed by an image displayed for 500ms. The image was either related to cannabis or neutral (52 cannabis/ 52 neutral). Following the image, a blank screen was displayed for 500ms, after which an arrow image, pointing either in the left or right direction, was presented. The participant was to respond to this arrow by pressing either a left or right button. The arrow image was released once the press was registered (maximum duration 1000ms). The arrow was followed by an interstimulus fixation cross with variable duration (900-2100ms, mean = 1477ms), which transitioned into the next trial's 200ms starting fixation cross. In

STOP trials, a stop signal was shown above the arrow following a delay, which indicated to the participant to inhibit the button press. The initial delay time was 250ms. Successful inhibition increased the time by 50ms while failed inhibition decreased the time by 50ms within a range of 0-500ms. Stop signal reaction times, taken as the inferred mean latency between the stop signal and response inhibition, were estimated via an additive multilevel linear model with a participant-varying intercept. See eFigure 4 for a graphical overview.

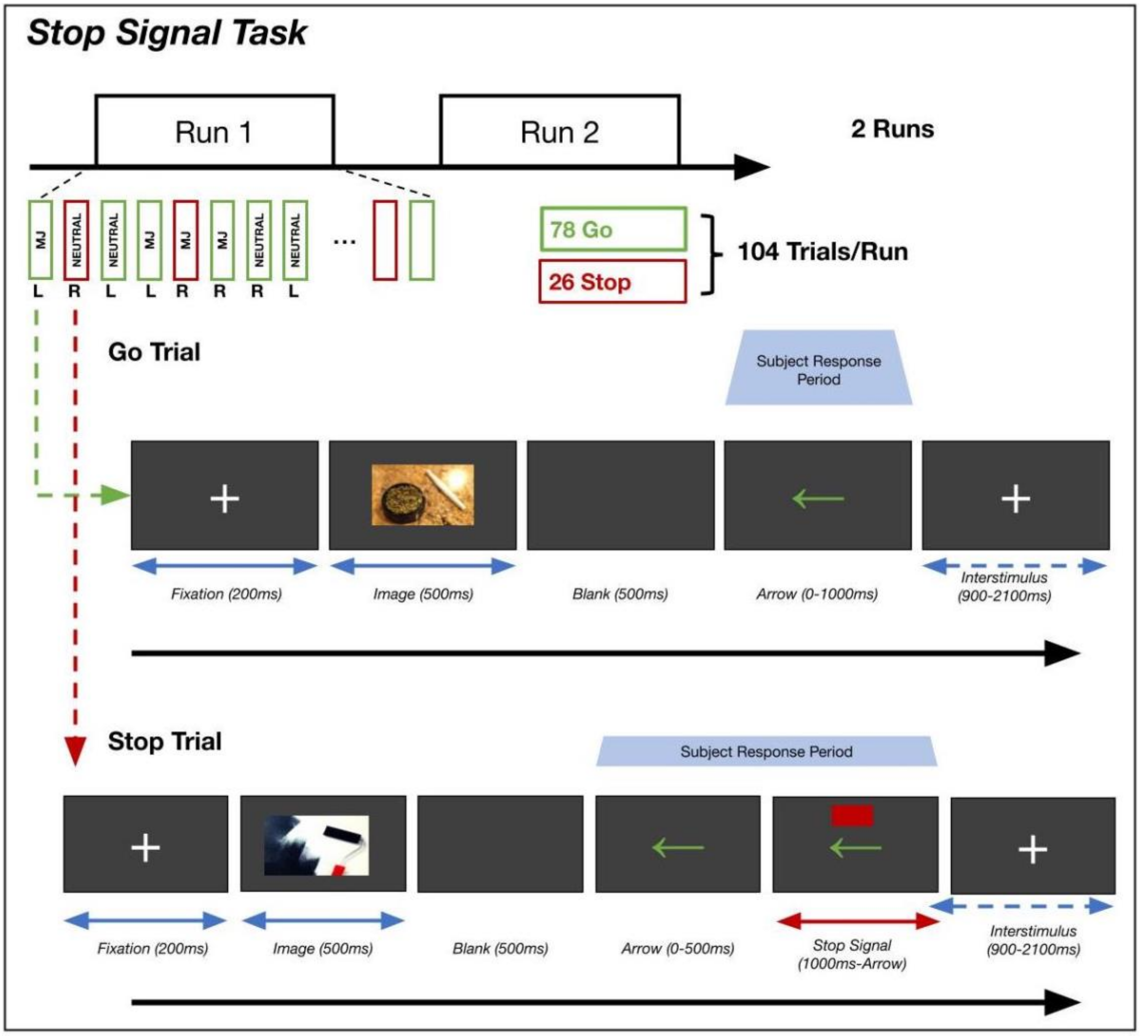


#### eFigure 4: Schematic of the SST task

In the SST task participants need to press a button as fast as possible when presented with an arrow unless a stop sign appears above the arrow with a slight delay. Some trials are preceded by a neutral and others by a cannabis-related image and there are two runs total.

##### MRI Data Analysis

###### MRI Acquisition

Stimuli were displayed on a MRI-compatible display system, which participants saw using a mirror attached to the head coil. MRI acquisitions were collected on a 3T Siemens Trio scanner with XQ Gradients (maximum amplitude of 45 mT/m and slew rate of 200 T/m/s) and a 32-channel head coil. Sagittal 3D T1-weighted images were collected using a MPRAGE sequence with a generalized autocalibrating partially parallel acquisition (GRAPPA) technique (repetition time (TR) = 2400ms, echo time (TE) = 2.02ms, inversion time (TI) = 1000ms, flip angle (FA) = 8°, slice thickness = 0.7 mm, field of view (FOV) = 256 × 256 mm, number of slices = 256, matrix size = 351 × 351, GRAPPA reduction factor = 2). T2*-weighted images were collected using an echo-planar imaging (EPI) sequence with a simultaneous multislice protocol (TR = 1530ms, TE = 30ms, FA = 80°, slice thickness = 2 mm, FOV = × mm, number of slices = 69, matrix size = 110 × 110). For the N-back, MID and SST tasks 278, 215 and 257 functional scans were acquired per run, respectively, and the total acquisition times were about 425, 329 and 393 seconds per run, respectively.

#### Pre-Processing

The following is explicitly copied according to the instructions from the *fMRIPrep* boilerplate generated during the *fMRIPrep 23.0.1* preprocessing. Changes made were the deletion of a duplicate paragraph and the changing of the citation style.

*Results included in this manuscript come from preprocessing performed using fMRIPrep 23.0.1 (RRID:SCR_016216),[1,2] which is based on Nipype 1.8.5 (RRID:SCR_002502).[3,4]*

***Preprocessing of B0 inhomogeneity mappings***

*A total of 1 fieldmaps were found available within the input BIDS structure for this particular subject. A B0-nonuniformity map (or fieldmap) was estimated based on two (or more) echo-planar imaging (EPI) references with topup (FSL 6.0.5.1:57b01774).[5]*

***Anatomical data preprocessing***

*A total of 1 T1-weighted (T1w) images were found within the input BIDS dataset.The T1-weighted (T1w) image was corrected for intensity non-uniformity (INU) with N4BiasFieldCorrection,[6] distributed with ANTs 2.3.3 (RRID:SCR_004757),[7] and used as T1w-reference throughout the workflow. The T1w-reference was then skull-stripped with a Nipype implementation of the antsBrainExtraction.sh workflow (from ANTs), using OASIS30ANTs as target template. Brain tissue segmentation of cerebrospinal fluid (CSF), white-matter (WM) and gray-matter (GM) was performed on the brain-extracted T1w using fast (FSL 6.0.5.1:57b01774, RRID:SCR_002823).[8] Brain surfaces were reconstructed using recon-all (FreeSurfer 7.3.2, RRID:SCR_001847),[9] and the brain mask estimated previously was refined with a custom variation of the method to reconcile ANTs-derived and FreeSurfer-derived segmentations of the cortical gray-matter of Mindboggle (RRID:SCR_002438).[10] Grayordinate "dscalar" files[11] containing 91k samples were also generated using the highest-resolution fsaverage as an intermediate standardized surface space. Volume-based spatial normalization to two standard spaces (MNI152NLin6Asym, MNI152NLin2009cAsym) was performed through nonlinear registration with antsRegistration (ANTs 2.3.3), using brain-extracted versions of both T1w reference and the T1w template. The following templates were were selected for spatial normalization and accessed with TemplateFlow (23.0.0):[12] FSL's MNI ICBM 152 non-linear 6th Generation Asymmetric Average Brain Stereotaxic Registration Model [RRID:SCR_002823; TemplateFlow ID: MNI152NLin6Asym],[13] ICBM 152 Nonlinear Asymmetrical template version 2009c [RRID:SCR_008796; TemplateFlow ID: MNI152NLin2009cAsym].[14]*

***Functional data preprocessing***

*For each of the 6 BOLD runs found per subject (across all tasks and sessions), the following preprocessing was performed. First, a reference volume and its skull-stripped version were generated using a custom methodology of fMRIPrep. Head-motion parameters with respect to the BOLD reference (transformation matrices, and six corresponding rotation and translation parameters) are estimated before any spatiotemporal filtering using mcflirt (FSL 6.0.5.1:57b01774).[15] The estimated fieldmap was then aligned with rigid-registration to the target EPI (echo-planar imaging) reference run. The field coefficients were mapped on to the reference EPI using the transform.*

*BOLD runs were slice-time corrected to 0.722s (0.5 of slice acquisition range 0s-1.45s) using 3dTshift from AFNI (RRID:SCR_005927).[16] The BOLD reference was then co-registered to the T1w reference using bbregister (FreeSurfer) which implements boundary-based registration.[17] Co-registration was configured with six degrees of freedom. Several confounding time-series were calculated based on the preprocessed BOLD: framewise displacement (FD), DVARS and three region-wise global signals. FD was computed using two formulations following Power (absolute sum of relative motions)[18] and Jenkinson (relative root mean square displacement between affines).[15] FD and DVARS are calculated for each functional run, both using their implementations in Nipype (following the definitions by Power et al. 2014). The three global signals are extracted within the CSF, the WM, and the whole-brain masks. Additionally, a set of physiological regressors were extracted to allow for component-based noise correction (CompCor).[19] Principal components are estimated after high-pass filtering the preprocessed BOLD time-series (using a discrete cosine filter with 128s cut-off) for the two CompCor variants: temporal (tCompCor) and anatomical (aCompCor). tCompCor components are then calculated from the top 2% variable voxels within the brain mask. For aCompCor, three probabilistic masks (CSF, WM and combined CSF+WM) are generated in anatomical space. The implementation differs from that of Behzadi et al. in that instead of eroding the masks by 2 pixels on BOLD space, a mask of pixels that likely contain a volume fraction of GM is subtracted from the aCompCor masks. This mask is obtained by dilating a GM mask extracted from the FreeSurfer's aseg segmentation, and it ensures components are not extracted from voxels containing a minimal fraction of GM. Finally, these masks are resampled into BOLD space and binarized by thresholding at 0.99 (as in the original implementation). Components are also calculated separately within the WM and CSF masks. For each CompCor decomposition, the k components with the largest singular values are retained, such that the retained components' time series are sufficient to explain 50 percent of variance across the nuisance mask (CSF, WM, combined, or temporal). The remaining components are dropped from consideration. The head-motion estimates calculated in the correction step were also placed within the corresponding confounds file. The confound time series derived from head motion estimates and global signals were expanded with the inclusion of temporal derivatives and quadratic terms for each.[20] Frames that exceeded a threshold of 0.5 mm FD or 1.5 standardized DVARS were annotated as motion outliers. Additional nuisance timeseries are calculated by means of principal components analysis of the signal found within a thin band (crown) of voxels around the edge of the brain, as proposed by.[21] The BOLD time-series were resampled into standard space, generating a preprocessed BOLD run in MNI152NLin6Asym space. First, a reference volume and its skull-stripped version were generated using a custom methodology of fMRIPrep. The BOLD time-series were resampled onto the following surfaces (FreeSurfer reconstruction nomenclature): fsaverage. Automatic removal of motion artifacts using independent component analysis (ICA-AROMA)[22] was performed on the preprocessed BOLD on MNI space time-series after removal of non-steady state volumes and spatial smoothing with an isotropic, Gaussian kernel of 6mm FWHM (full-width half-maximum). Corresponding "non-aggresively" denoised runs were produced after such smoothing. Additionally, the "aggressive" noise-regressors were collected and placed in the corresponding confounds file. Grayordinates files[11] containing 91k samples were also generated using the highest-resolution fsaverage as intermediate standardized surface space. All resamplings can be performed with a single interpolation step by composing all the pertinent transformations (i.e. head-motion transform matrices, susceptibility distortion correction when available, and co-registrations to anatomical and output spaces). Gridded (volumetric) resamplings were performed using antsApplyTransforms (ANTs), configured with Lanczos interpolation to minimize the smoothing effects of other kernels.[23] Non-gridded (surface) resamplings were performed using mri_vol2surf (FreeSurfer).*

*Many internal operations of fMRIPrep use Nilearn 0.9.1 (RRID:SCR_001362),[24] mostly within the functional processing workflow. For more details of the pipeline, see the section corresponding to workflows in fMRIPrep's documentation.*

##### First-Level Analysis

Data in grayordinate format used in the surface-based analysis was smoothed using the Human Connectome Project (HCP) workbench toolbox with the surface and volume kernel both set to 1.6986 (corresponds to a full width at half maximum (fwhm) of 4mm). Smoothing of voxels for the volume-based analysis (fwhm set to 4mm) and general linear model (GLM) fitting were conducted using the Python package *Nilearn* version 0.9.2. Post smoothing and pre GLM, the time series was scaled by multiplying by a constant, 1000, and dividing by the median of the grayordinates across time and space, after subtracting the minimum value of the time series across time and space from both the time series and the median. First-level modeling using a multiple linear model included as nuisance regressors six rigid head motion parameters, single-volume motion and non-steady state outliers, discrete cosine-basis regressors, the first six anatomical CompCor regressors and framewise displacement. Noise was modeled using a first-order autoregressive model. Stimuli regressors used in the first-level modeling included regressors for the zero-back and the two-back blocks of the N-back task, cue, hit and miss regressors for the high/low reward, high/low loss, and neutral events of the MID task, and marijuana and neutral-primed regressors for the GO, successful STOP, and unsuccessful GO events of the SST task. These were convolved with a hemodynamic response function from the SPM dispersion derivative model. For the N-back task, the contrast was two-back vs. zero-back. For the MID task, anticipation contrasts included high reward vs. neutral anticipation, low reward vs. neutral anticipation, reward vs. neutral anticipation, high reward vs. low reward anticipation, high reward vs. implicit baseline, high loss vs. neutral anticipation, low loss vs. neutral anticipation, and high loss vs. low loss anticipation. Further, feedback contrasts included high reward vs. neutral hit feedback, reward vs. missed reward feedback, high loss vs. neutral hit feedback and loss vs. avoided loss feedback. For the SST task, the contrasts were correct inhibition (successful STOP vs. GO trials), incorrect inhibition (unsuccessful STOP vs. GO trials), and unsuccessful inhibitory control (unsuccessful STOP vs. successful STOP trials). For the MID and the SST tasks, the contrasts from the two runs were combined using a second linear model prior to group-level modeling.

##### Group-Level Analysis

The effect sizes of group contrasts were taken to be the average (for group level results at a single time point), average difference (for differences in the same participants across time) and difference of averages (for differences across the control and MCC participant groups at baseline) of the individual contrasts of the participants. These were estimated using an ordinary least squares multiple regression model and significance of the group contrasts was assessed in a two-sided manner. Imaging results were FDR controlled at 0.05. Effect sizes at the group level were standardized for visualization purposes using the unbiased Hedges estimator. Covariates in the group-level model included sex, age, and past-month cannabis use frequency, mean-centered for numerical variables. Of note, past-month cannabis use frequency, originally collected as a categorical variable, was re-coded as a numerical variable representing the approximate fraction of days in a month where cannabis was used. In the supplemental sensitivity analysis of using an FD cutoff of 0.3 and no outlier removal, an additional subject had to be excluded due to a missing past-month cannabis use frequency value at baseline.

#### Supplementary Results

##### Cannabis Metrics

| Items | Levels | MCC baseline[a] | MCC one-year[b] |
|---|---|---|---|
| CUD[c] diagnosis, n (%) | | 0 | 6 (14.6)[d] |
| CUDIT[e] summed score, mean (SD) | | 2.4 (2.5) | 4.8 (2.4) |
| THC[f] frequency per month, n (%) | Less than once a month | 27 (65.9) | 6 (14.6) |
| THC frequency per month, n (%) | Less than once every two weeks | 2 (4.9) | 2 (4.9) |
| THC frequency per month, n (%) | Less than once a week | 6 (14.6) | 4 (9.8) |
| THC frequency per month, n (%) | 1-2 days a week | 2 (4.9) | 9 (22.0) |
| THC frequency per month, n (%) | 3-4 days a week | 4 (9.8) | 9 (22.0) |
| THC frequency per month, n (%) | 5-6 days a week | 0 | 3 (7.3) |
| THC frequency per month, n (%) | Once or more per day | 0 | 8 (19.5) |
| Positive urine THC, n (%) | | 5 (12.2) | 15 (38.5) |

**eTable 1: Statistically significant difference in cannabis-related characteristics after one year of cannabis use for medical symptoms**

[a] MCC baseline corresponds to the participants of the medical cannabis group, who had imaging collected at both timepoints, at baseline

[b] MCC one-year corresponds to the participants of the medical cannabis group, who had imaging collected at both timepoints, at baseline

[c] CUD = cannabis use disorder

[d] All CUD diagnoses were mild (2-3 symptoms)

[e] CUDIT = Cannabis Use Disorder Identification Test

[f] THC = tetrahidrocannabinol.

N-back Accuracy and Reaction Time

| | HC vs. MCC at baseline[a] | | MCC at baseline vs. one-year[b] | |
|---|---|---|---|---|
| | difference of means | p value | mean difference | p value |
| Combined accuracy | -0.001 | 0.989 | 0.018 | 0.656 |
| Combined reaction time | 6.279 | 0.652 | -4.8 | 0.712 |
| Accuracy of zero-back trials | -0.008 | 0.877 | 0.027 | 0.602 |
| Reaction time of zero-back trials | 1.846 | 0.875 | -5.899 | 0.654 |
| Accuracy of two-back trials | 0.006 | 0.878 | 0.009 | 0.789 |
| Reaction time of two-back trials | 10.604 | 0.606 | -1.576 | 0.918 |

**eTable 2: No statistically significant difference in N-back accuracy and reaction time compared to control or across time**

[a] HC vs. MCC at baseline corresponds to comparing the imaging control group with the medical cannabis group at baseline
[b] MCC at baseline vs. one-year corresponds to comparing the participants of the medical cannabis group with imaging at both timepoints across time

##### Results From Additional Contrasts

The following results are showing contrasts of the MID and SST tasks in addition to those in the main paper.

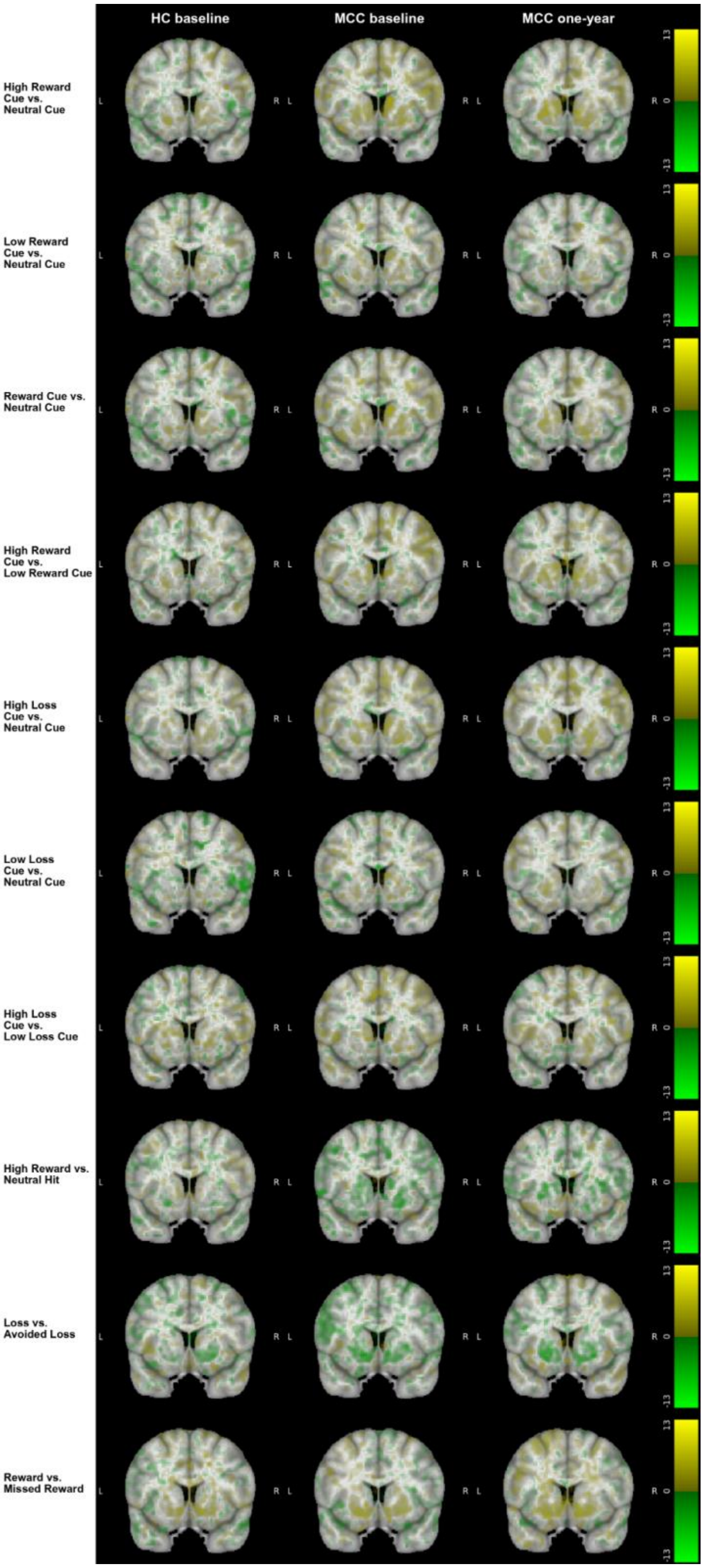

**eFigure 5: Brain activation for additional contrasts of the MID task across groups and timepoints from the volumetric analysis**

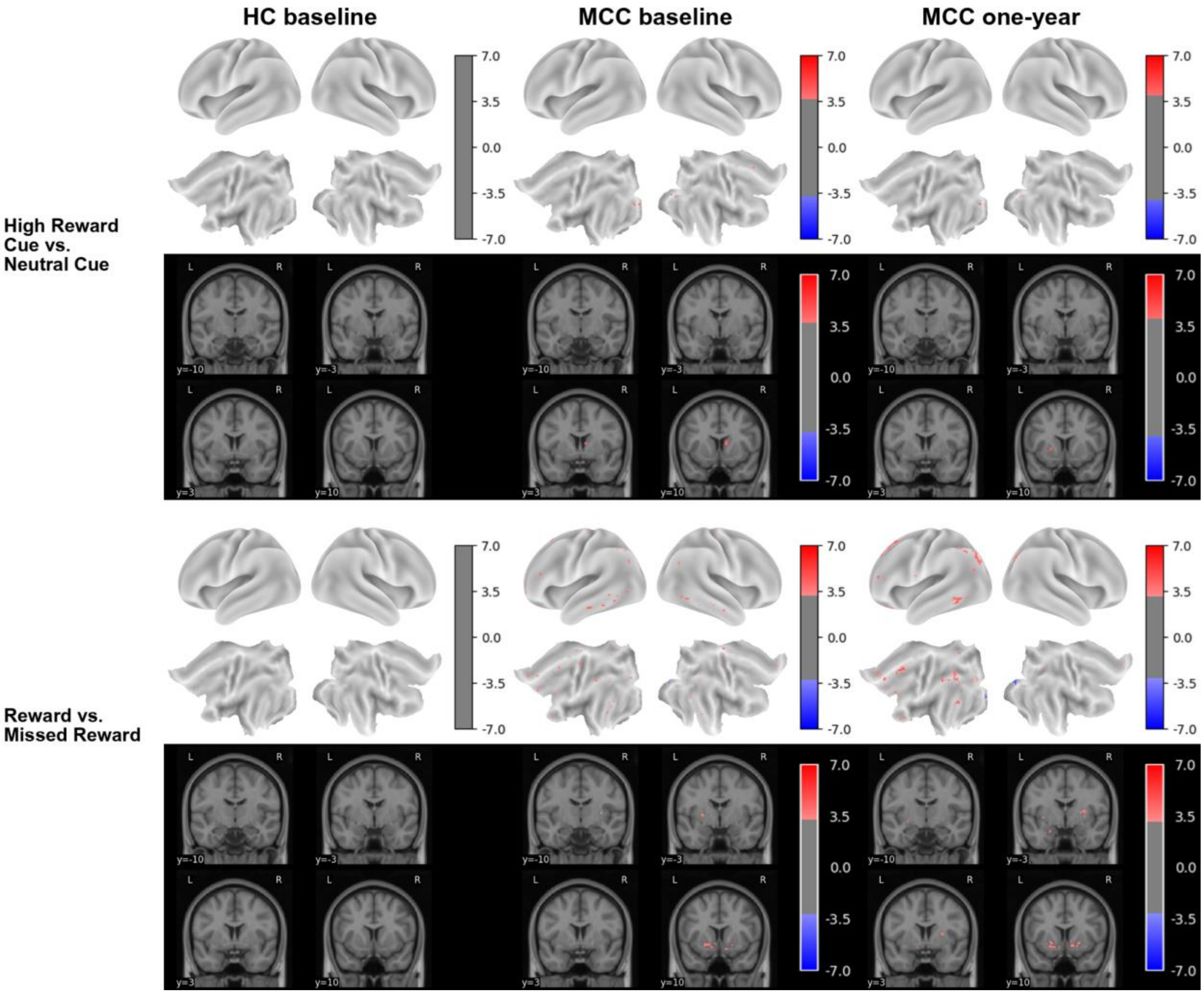


**eFigure 6: Brain activation for additional contrasts of the MID task across groups and timepoints from the grayordinate analysis**

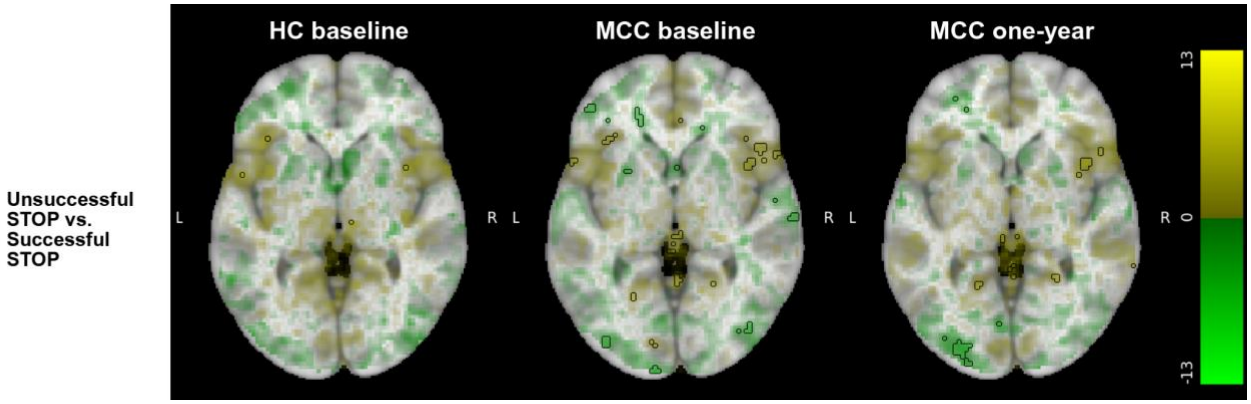


eFigure 7: Brain activation for an additional contrast of the SST task across groups and timepoints from the volumetric analysis

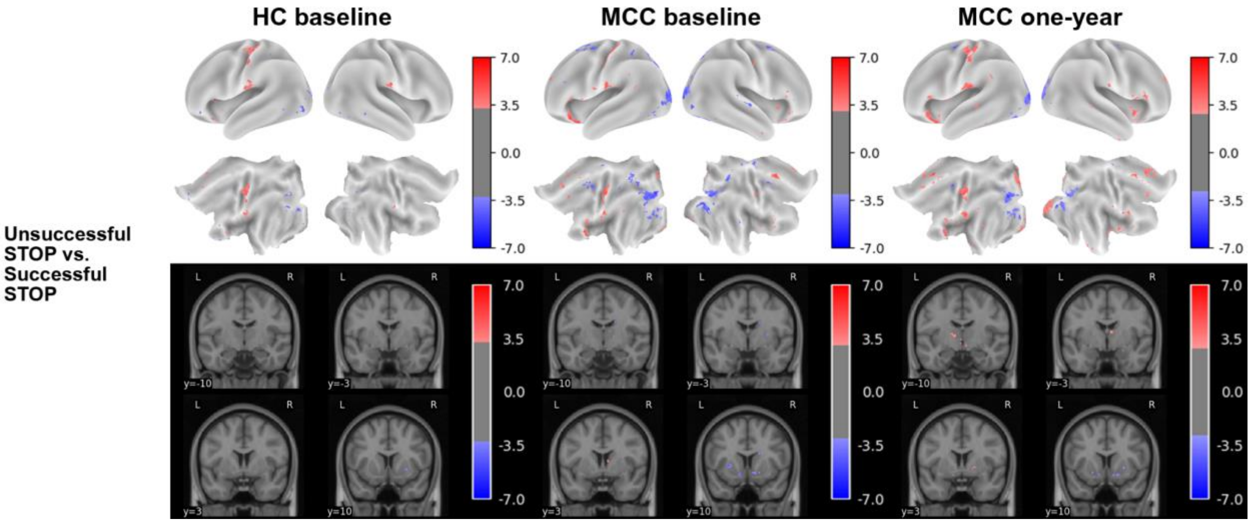


eFigure 8: Brain activation for an additional contrast of the SST task across groups and timepoints from the grayordinate analysis

#### Main Results With Varying Outlier Removal

The following results are showing the same contrasts as in the main paper but the scans removed due to being quality control outliers are varied. Shown below are the results when no outlier scans are removed and when the framewise displacement (FD) cutoff of 0.2 from the main results is relaxed to 0.3.

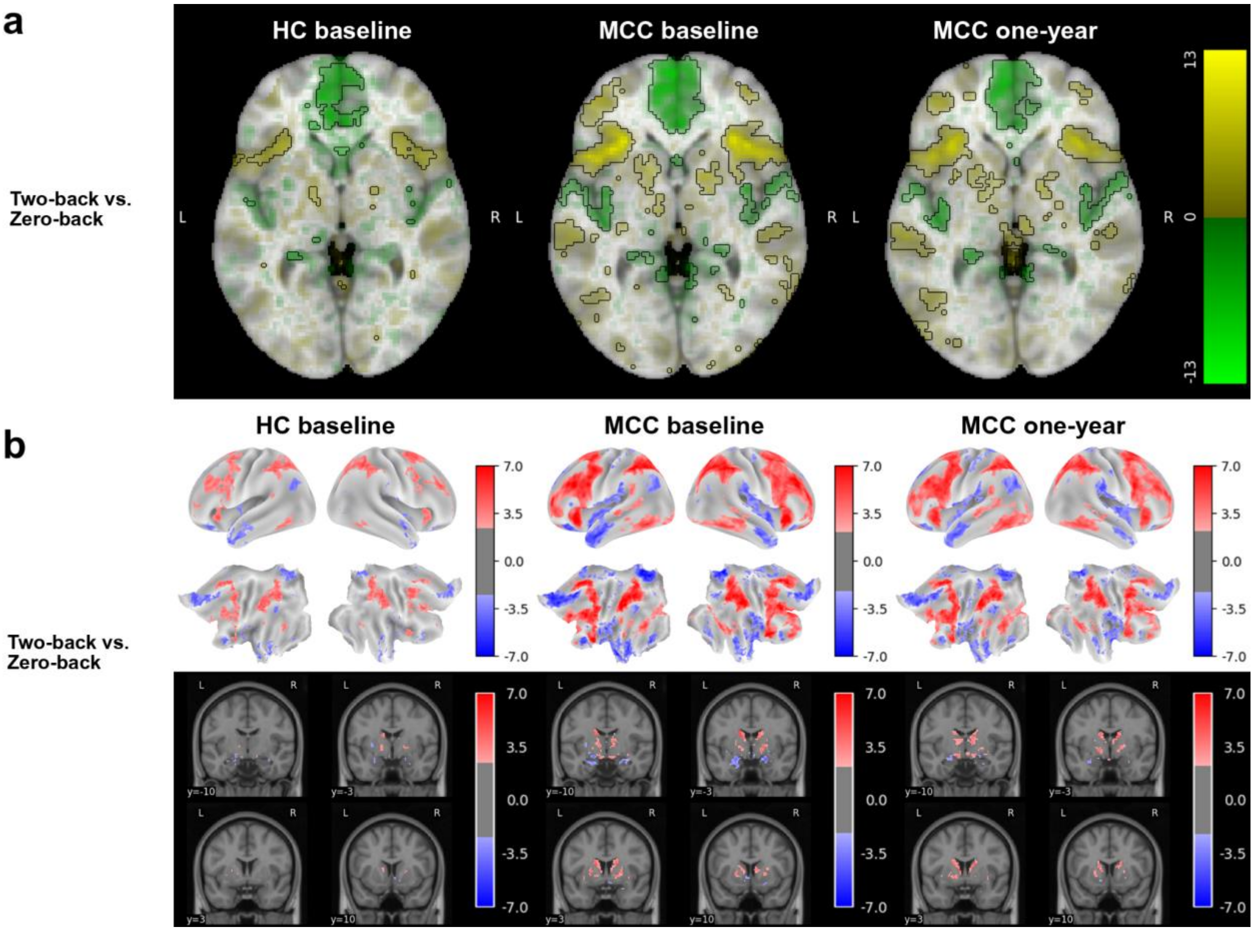


**eFigure 9: Brain activation for the N-back task's two-back vs. zero-back contrast across groups and timepoints with no outliers removed**

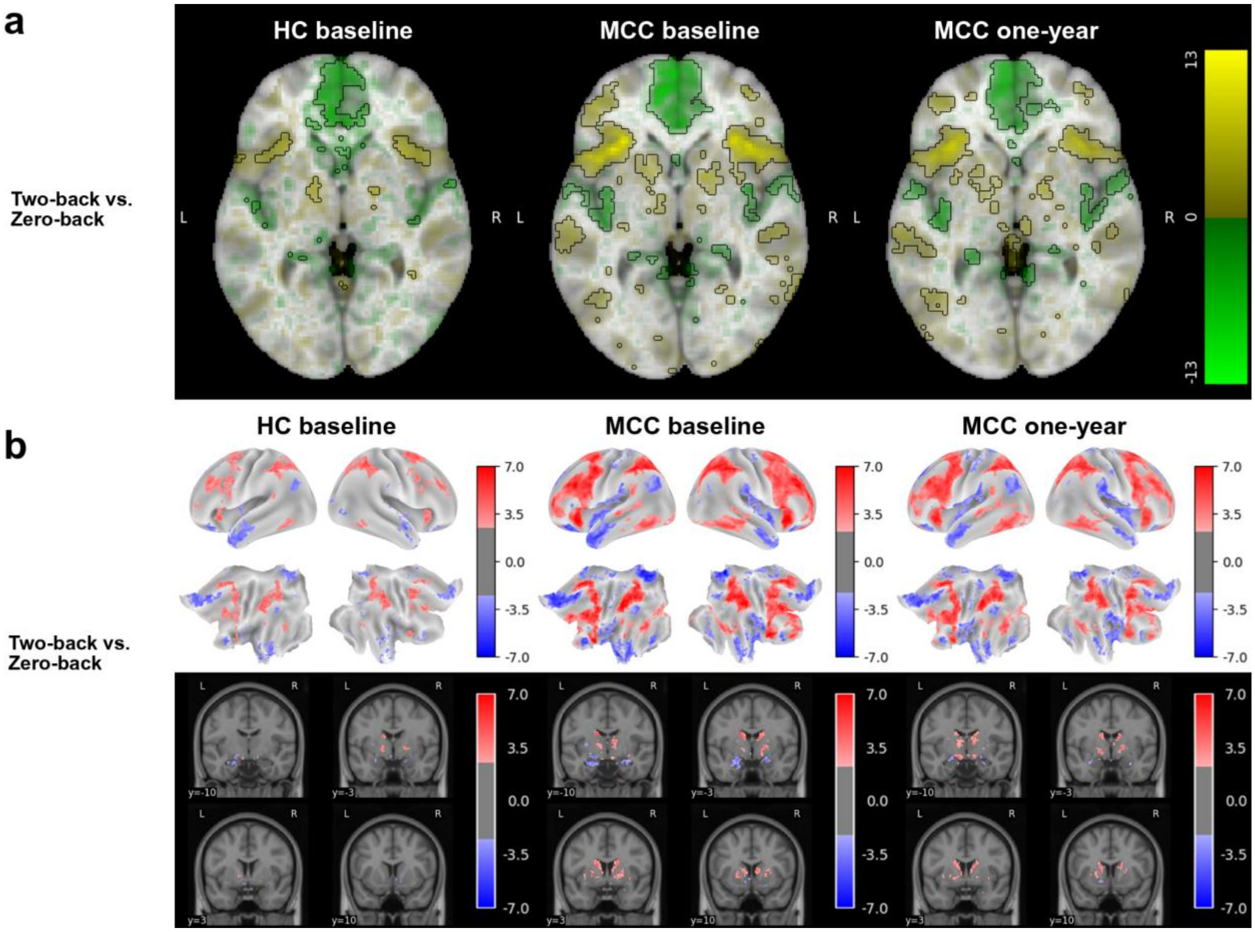


**eFigure 10: Brain activation for the N-back task's two-back vs. zero-back contrast across groups and timepoints with the FD cutoff relaxed to 0.3**

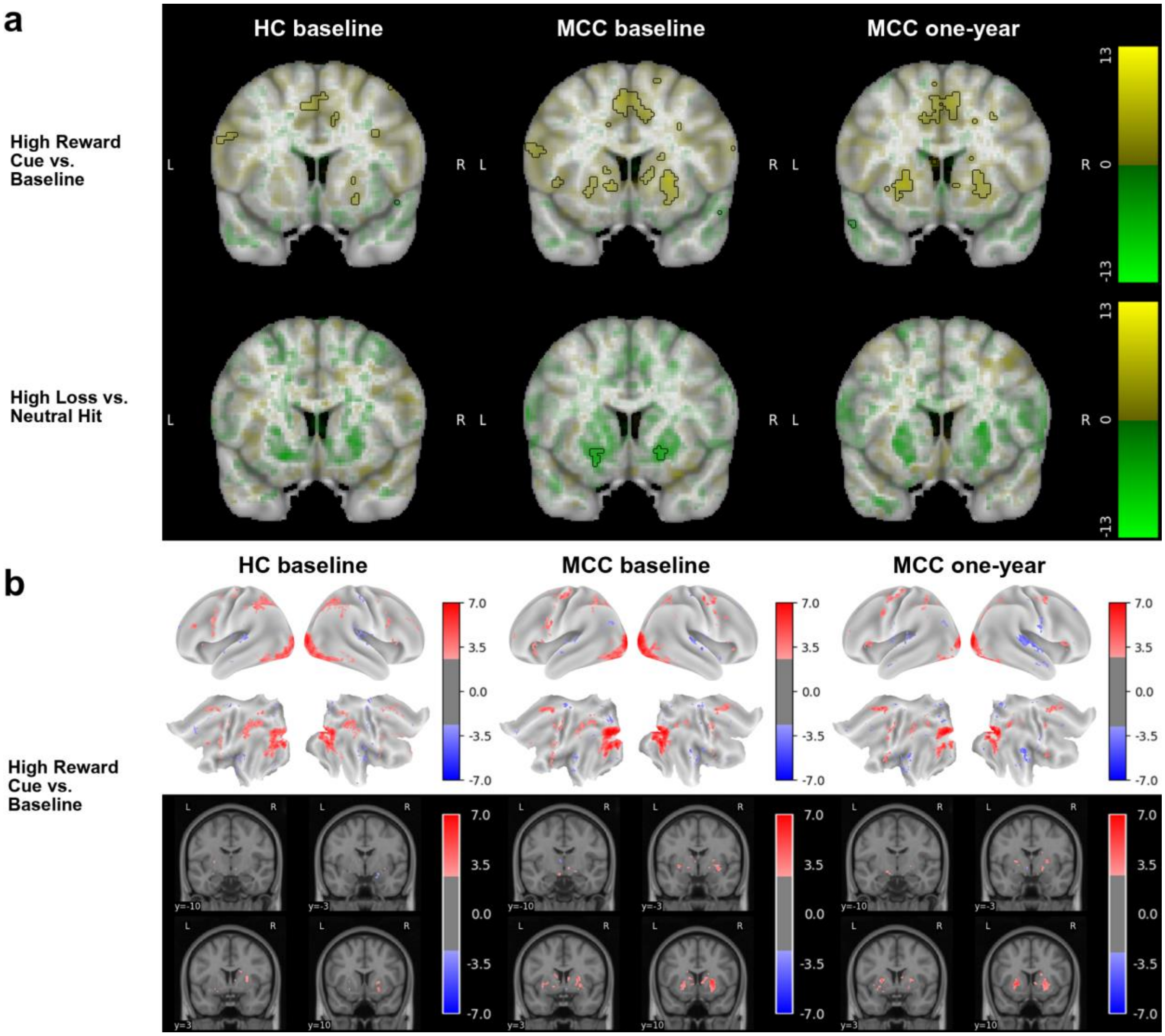


**eFigure 11: Brain activation for various contrasts of the MID task across groups and timepoints with no outliers removed**

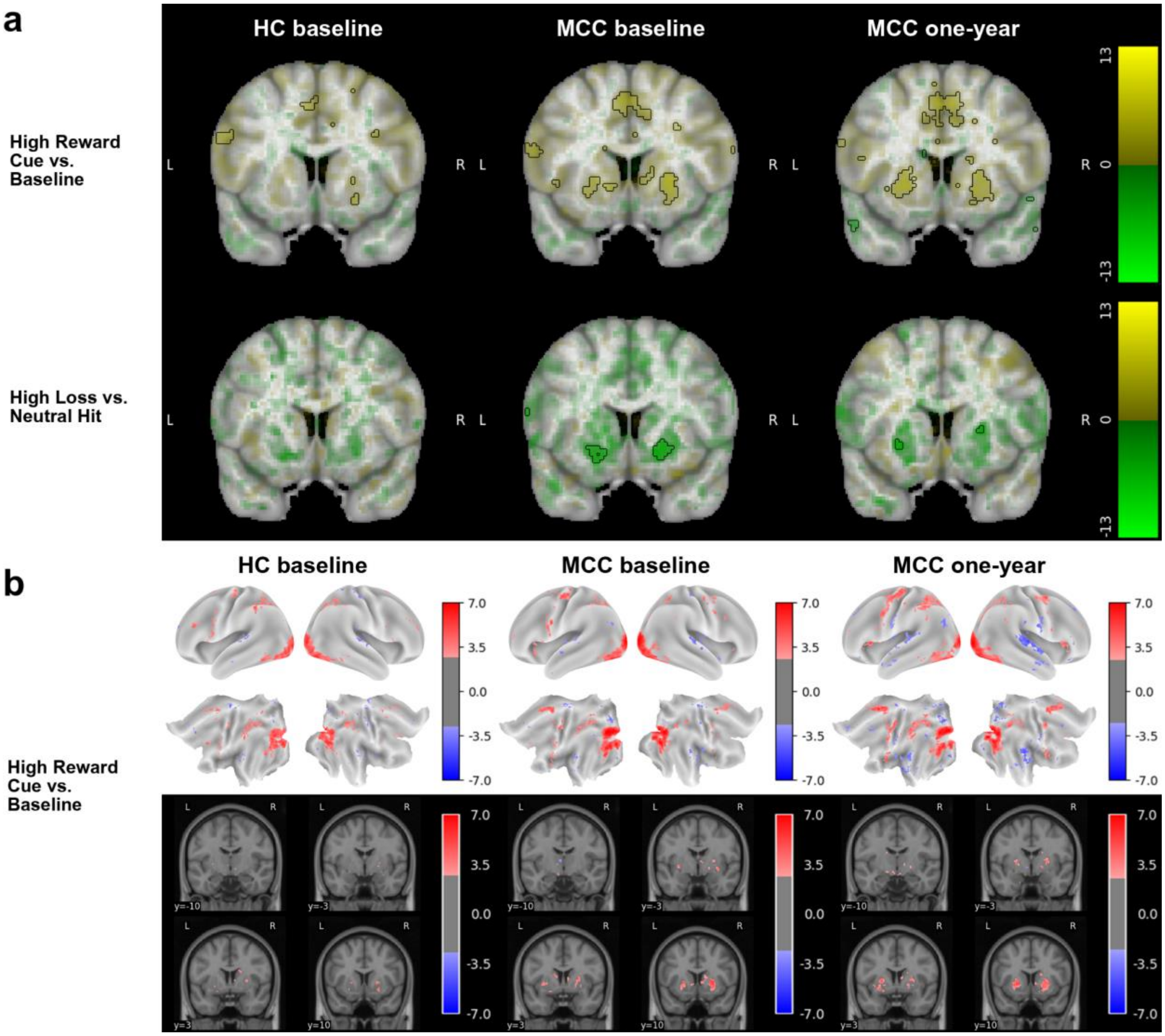


**eFigure 12: Brain activation for various contrasts of the MID task across groups and timepoints with the FD cutoff relaxed to 0.3**

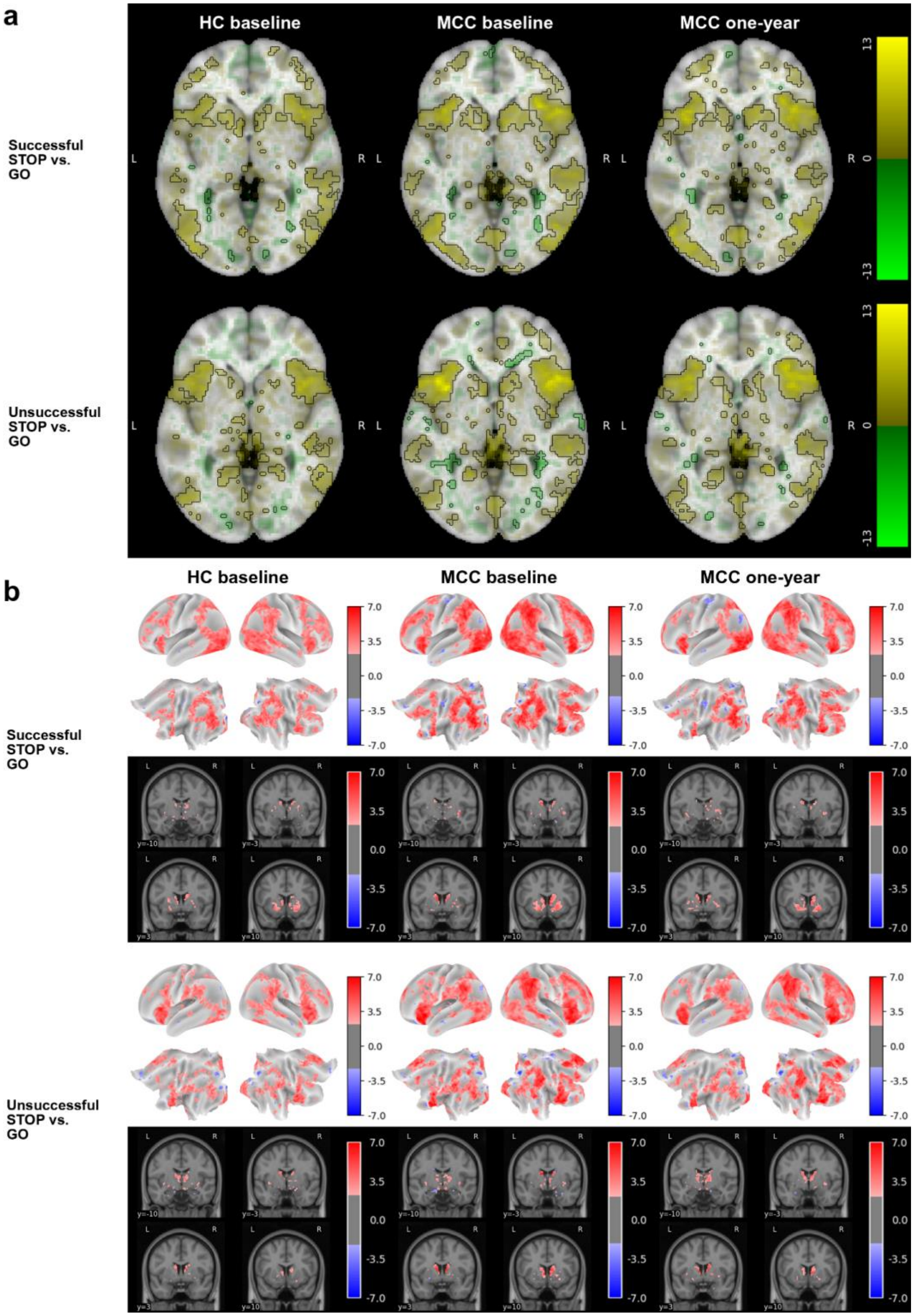


**eFigure 13: Brain activation for the SST task's two STOP vs. GO contrasts across groups and timepoints with no outliers removed**

a

b

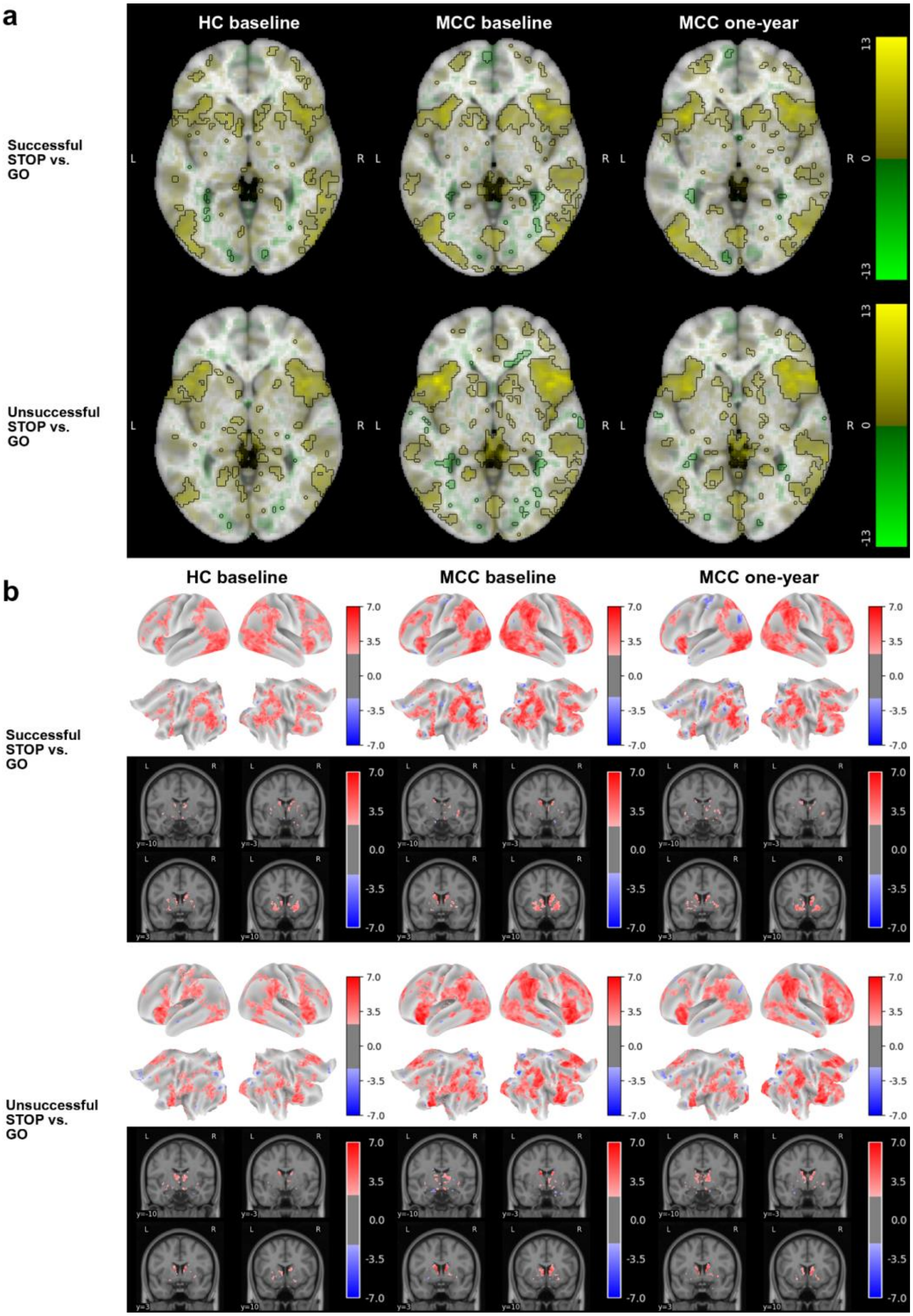


eFigure 14: Brain activation for the SST task's two STOP vs. GO contrasts across groups and timepoints with the FD cutoff relaxed to 0.3
